# Mortality from injury, overdose and suicide during the 2020 COVID-19 pandemic, March-July, 2020

**DOI:** 10.1101/2021.02.13.21251682

**Authors:** Jeremy S. Faust, Chengan Du, Katherine Dickerson Mayes, Shu-Xia Li, Zhenqiu Lin, Michael L. Barnett, Harlan M. Krumholz

**Affiliations:** Harvard Medical School, Brigham and Women’s Hospital Department of Emergency Medicine, Division of Health Policy and Public Health; Yale University School of Medicine, Yale New Haven Hospital, Center for Outcomes Research and Evaluation, New Haven, CT; Harvard Affiliated Emergency Medicine Residency, Boston, Massachusetts; Yale New Haven Hospital, Center for Outcomes Research and Evaluation New Haven, CT; Department of Health Policy and Management, Harvard T. H. Chan School of Public Health, Boston, MA

## Abstract

**Introduction**

The COVID-19 pandemic has been associated with substantial rates of all-cause excess mortality. The contribution of external causes of death to excess mortality including drug overdose, homicide, suicide, and unintentional injuries during the initial outbreak in the United States is less well documented.

**Methods:** Using public data published by the National Center for Health Statistics on February 10, 2021, we measured monthly excess mortality (the gap between observed and expected deaths) from five external causes using national-level data published by National Center for Health Statistics; assault (homicide); intentional self-harm (suicide); accidents (unintentional injuries); and motor vehicle accidents. We used seasonal autoregressive integrated moving average (sARIMA) models developed with cause-specific monthly mortality counts and US population data from 2015-2019 and estimated the contribution of individual cause-specific mortality to all-cause excess mortality from March-July 2020.

**Results:** From March-July, 2020, 212,825 (95% CI 136,236-290,776) all-cause excess deaths occurred in the US). There were 8,540 excess drug overdoses (all intents) (95% CI 5,106 to 11,975), accounting for 4% of all excess mortality; 1,455 excess homicide deaths (95% CI 708 to 2202, accounting for 0.7% of excess mortality; 5,492 excess deaths due to unintentional accidents occurred (95% CI 85 to 10,899, accounting for 2.6% of excess mortality. Though a non-significantly 135 (95% CI -1361 to 1,630) more MVA deaths were recorded during the study period, a significant decrease in April (525; 95% CI -817 to -233) and significant increases in June-July (965; 95% CI 348 to 1,587) were observed. Suicide deaths were statistically lower than projected by 2,067 (95% CI 941-3,193 fewer deaths).

**Meaning:** Excess deaths from drug overdoses, homicide, and addicents occurred during the pandemic but represented a small fraction of all-cause excess mortality. The excess external causes of death, however, still represent thousands of lives lost. Notably, deaths from suicide were lower than expected and therefore did not contribute to excess mortality.

## Introduction

All-cause excess mortality has been closely tracked during the COVID-19 pandemic. However, the contribution of external causes of death during the initial months of the US outbreak is incompletely understood.^1–3^ We analyzed monthly trends in drug overdose deaths, homicide, suicide, unintentional injuries, and motor vehicle accidents (MVAs) from 2015-2020, focusing on the first five months of the COVID-19 outbreak.

## Methods

We measured monthly excess mortality (the gap between observed and expected deaths) from five external causes using national-level data published by National Center for Health Statistics (NCHS) including provisional mortality figures through July 2020 (released February 2021). Data from March-July, 2020 was aggregated by the NCHS into five groups: drug overdose (all intents), *International Classification of Diseases, Version-10* codes X40-44, X60-64, X85, Y10-Y14; assault (homicide), U01-U02, X85-Y09, Y87.1); intentional self-harm (suicide). U03, X60-X84, Y87.0; accidents (unintentional injuries), V01-X59, Y85-Y86); motor vehicle accidents, V02– V04,V09.0,V09.2,V12–V14,V19.0–V19.2,V19.4–V19.6, V20–V79,V80.3–V80.5, V81.0– V81.1,V82.0–V82.1,V83–V86, V87.0–V87.8,V88.0–V88.8, V89.0,V89.2.^4,5^

To forecast all-cause and cause-specific expected monthly deaths from March-July, 2020, we used seasonal autoregressive integrated moving average (sARIMA) models developed with cause-specific monthly mortality counts and US population data from 2015-2019. We used the Akaike information criterion to select the best models. We plotted observed and expected deaths monthly with 95% CI estimated from sARIMA models. We estimated the contribution of individual cause-specific mortality to all-cause excess mortality by dividing cause-specific mortality by total all-cause excess mortality from March-July 2020.

This study used publicly available data and was not subject to institutional review approval and adhered to STROBE guidelines.

## Results

From March-July, 2020, 212,825 (95% CI 136,236-290,776) all-cause excess deaths occurred in the US (1,409,826 observed; 1,197,001 expected). There were 8,540 excess drug overdoses (all intents) (95% CI 5,106-11,975, Figure 1a), accounting for 4% of all excess mortality; 1,455 excess homicide deaths (95% CI 708-2202, Figure 1b), accounting for 0.7% of excess mortality; 5,492 excess deaths due to unintentional accidents occurred (95% CI 85-10,899, Figure 1c), accounting for 2.6% of excess mortality. Though a non-significantly 135 (95% CI -1361-1,630) more MVA deaths were recorded during the study period, a significant decrease in April (525; 95% CI -817 to - 233) and significant increases in June-July (965; 95% CI 348-1,587) were observed (Figure 1d). Suicide deaths were statistically lower than projected by 2,067 (95% CI 941-3,193 fewer deaths), (Figure 1e).

**Fig 1.**
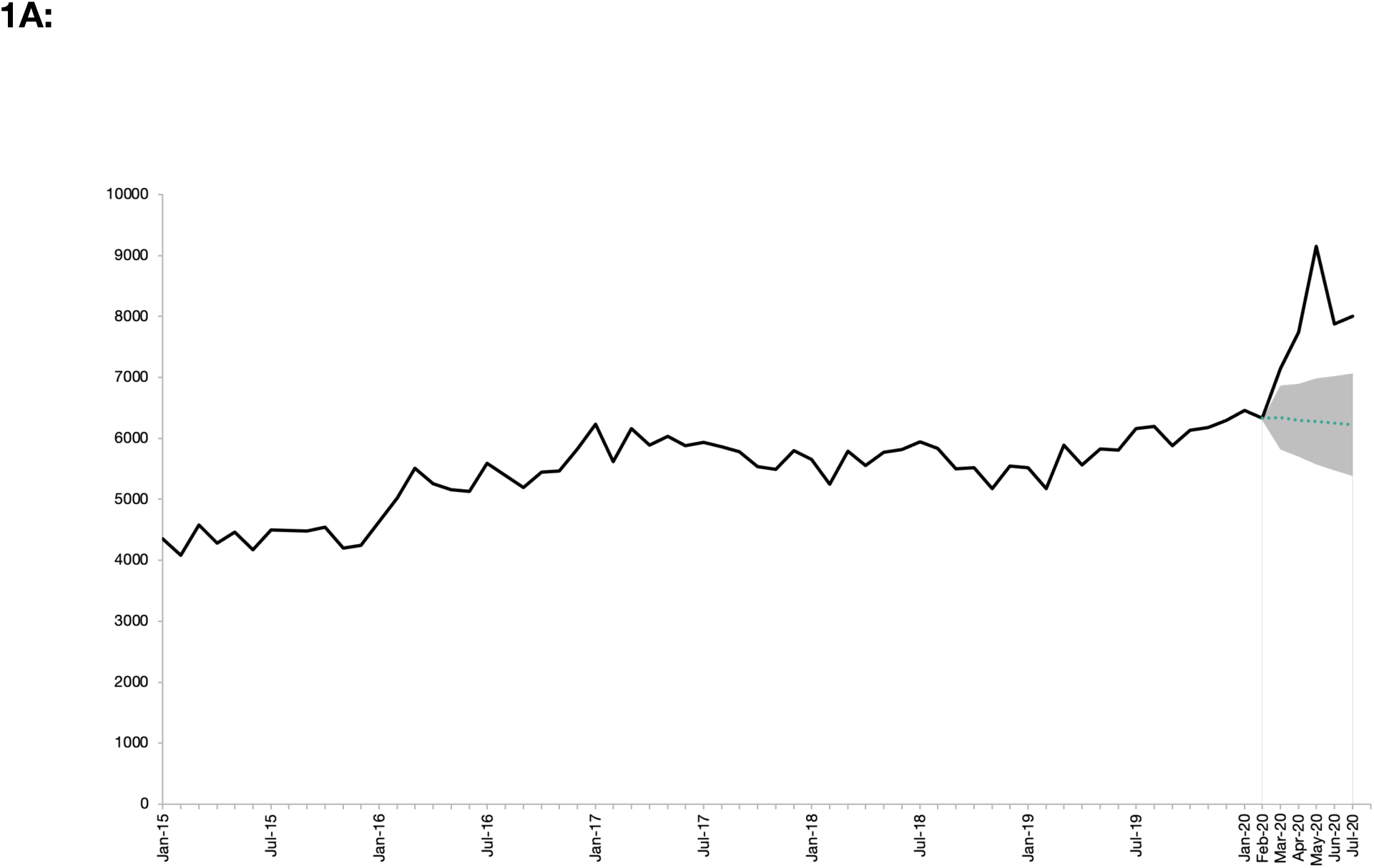
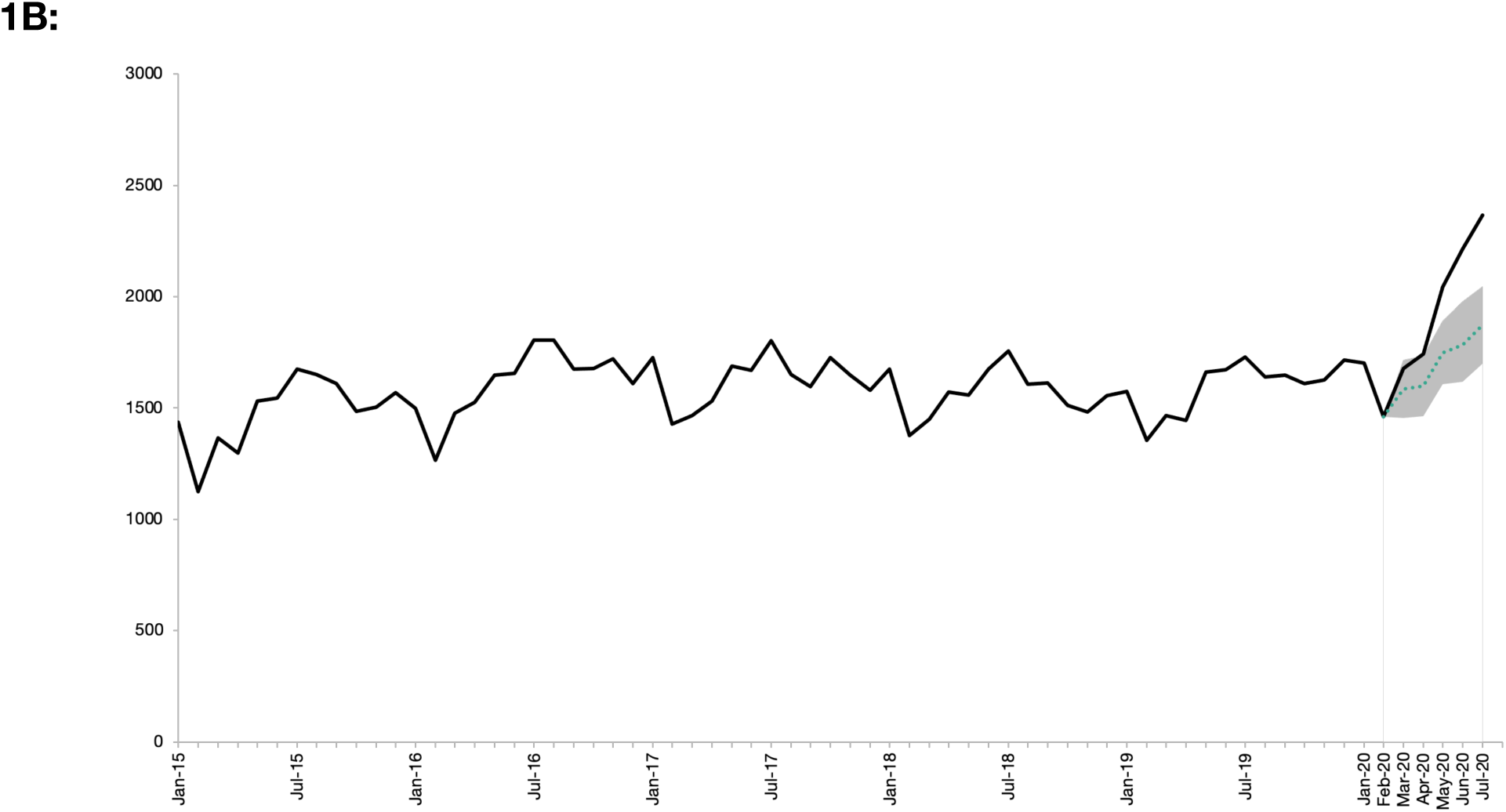
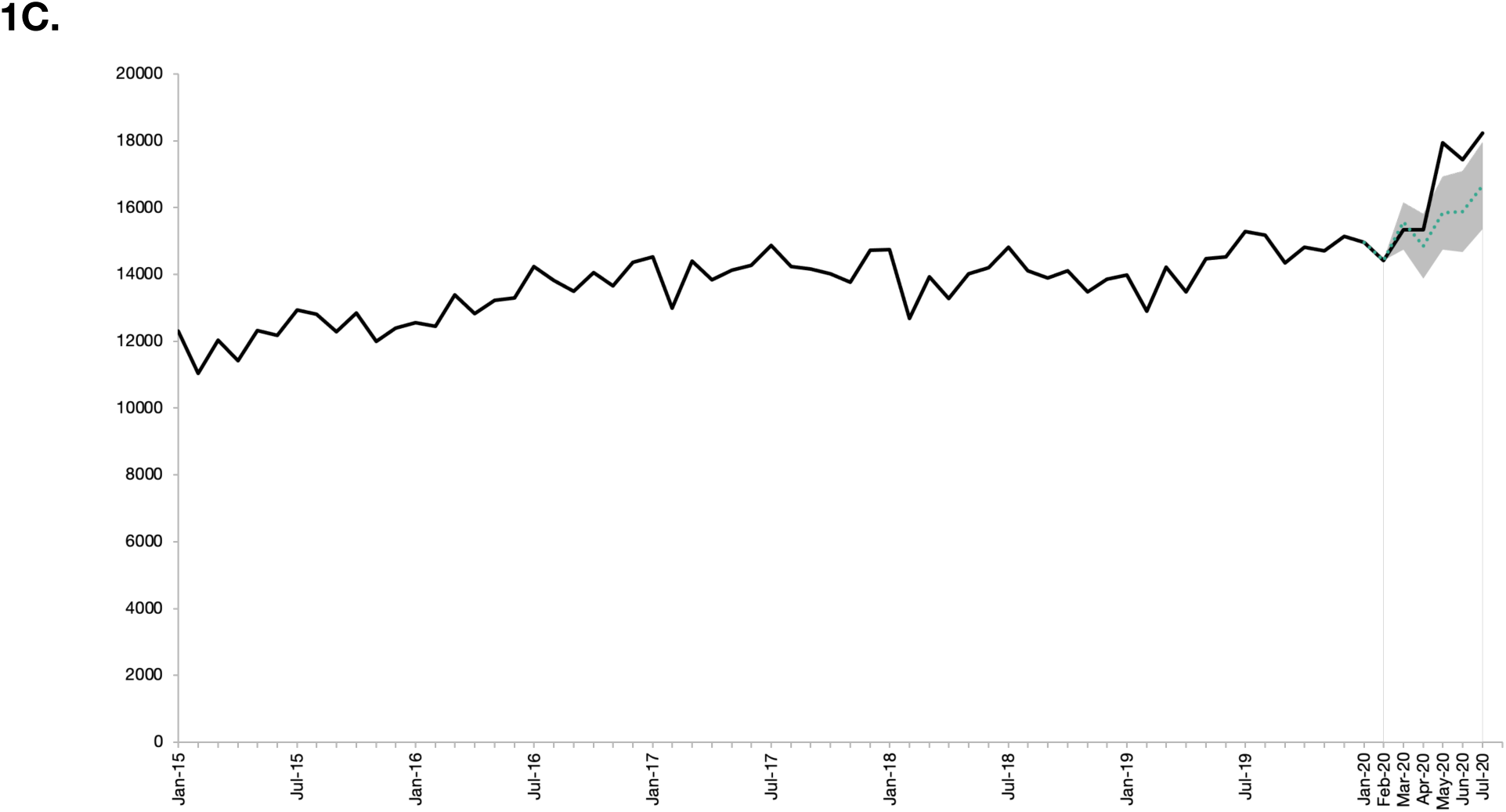
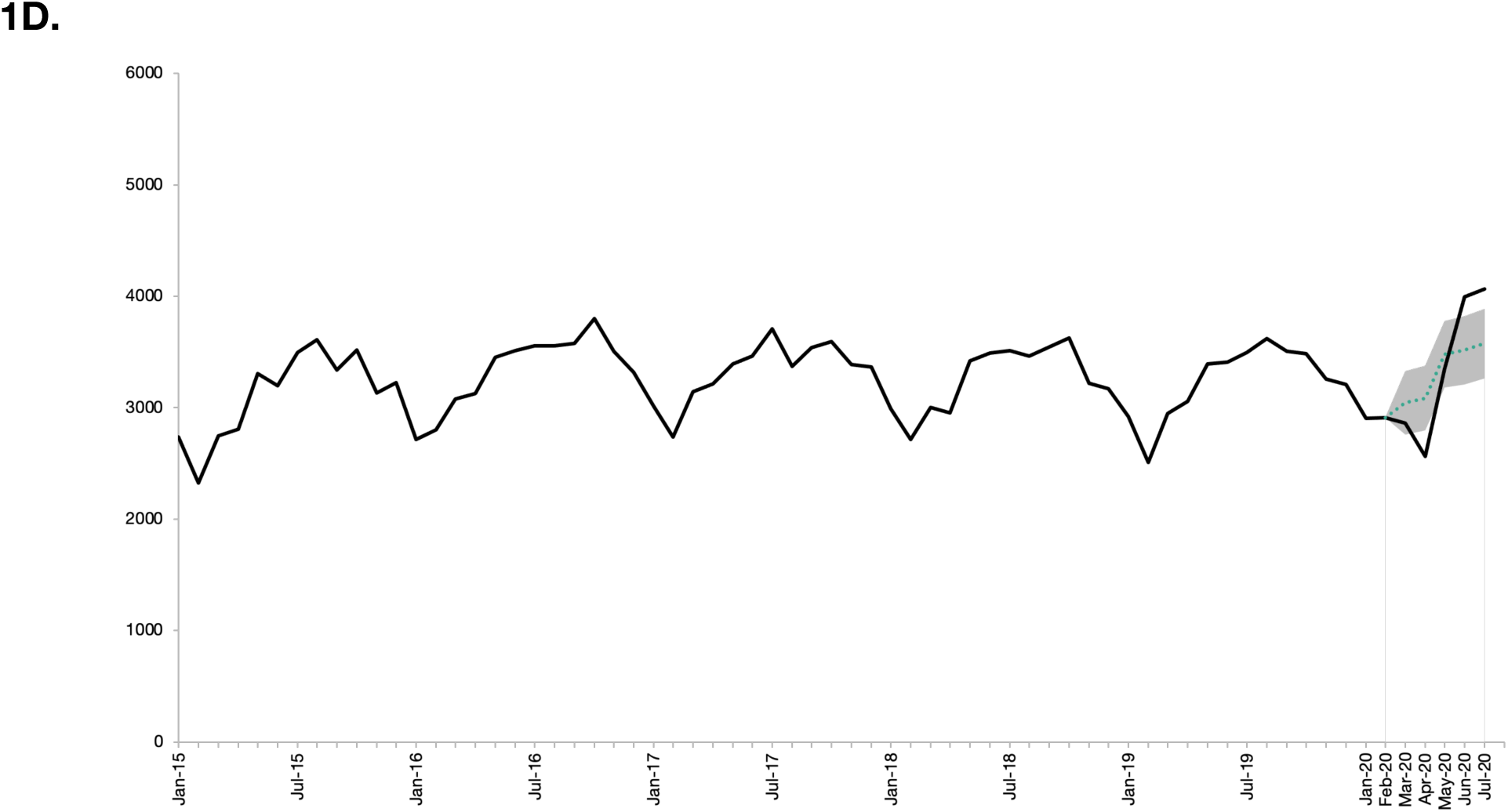
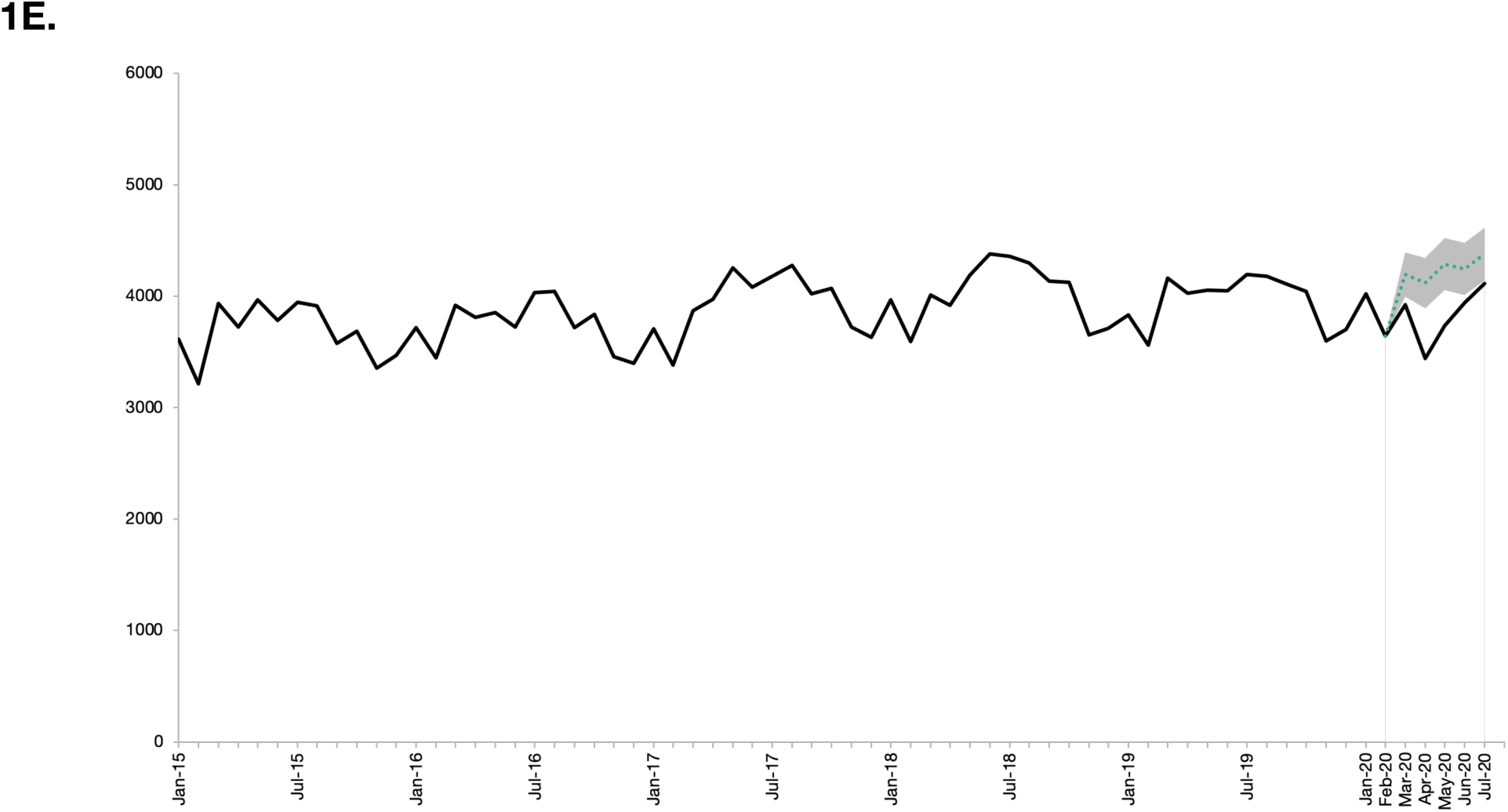
**Figure Title:** Cause-specific mortality in the United States, January 2015 through July 2020. **Figure Legend:** The solid black line indicates raw cause-specific death counts from January 2015-July 2020; the dashed green line represents the point estimate for cause-specific expected deaths from March-July 2020 using the seasonal adjusted model; the gray shaded area represents projected range (95% CI) of cause-specific deaths expected to occur during March-July 2020 using the seasonal adjusted model. 1A. Drug overdose deaths (all intents), United States, January 1, 2015 – July 31^st^, 2020. 1B. Homicide deaths, United States, January 1, 2015 – July 31^st^, 2020. 1C. Unintentional accident deaths, United States, January 1, 2015 – July 31^st^, 2020. 1D. Motor vehicle accident deaths, United States, January 1, 2015 – July 31^st^, 2020. 1E. Suicide deaths, United States, January 1, 2015 – July 31^st^, 2020.

## Discussion

Using provisional mortality data, we find that excess deaths from external causes occurred during the pandemic but represented a small fraction of all-cause excess mortality. The excess external causes of death, however, still represent thousands of lives lost exceeding pre-pandemic trends. Notably, deaths from suicide were lower than expected and therefore did not contribute to excess mortality.

Explanations for these excess external deaths are uncertain. Drug overdoses and homicides may be related to economic stress. Pandemic-associated changes in access to substance use disorder treatments could be exacerbating mortality from overdoses.^6^ Decreases in MVA deaths in April coincided with less traffic, despite increases in drivers testing positive for drugs and alcohol and lower seatbelt use.^3^ Increases in MVA deaths in June-July occurred as traffic increased (though still below 2019 levels), likely reflecting higher-risk behaviors.^3^

A limitation of this study is that all 2020 data published by NCHS are considered preliminary. However, substantial changes to March-July 2020 data are unlikely.

The increases in external causes of death may have been preventable. These findings suggest the need for public health strategies, both now and in future outbreaks, to include attention on these and other socio-behavioral risks. These data suggest that enhanced safety measures may prove beneficial, not just during stay-at-home periods but during subsequent re-opening phases as well.

## Data Availability

Dr. Faust had full access to all of the data in the study and takes responsibility for the integrity of the data and the accuracy of the data analysis. The data are available to the public.

## Data Statement and Author Contributions

Dr. Faust had full access to all of the data in the study and takes responsibility for the integrity of the data and the accuracy of the data analysis.

*Concept and design:* Faust, Barnett, Krumholz

*Acquisition, analysis, or interpretation of data:* Faust, Du, Mayes, Li, Lin

*Drafting of the manuscript:* Faust

*Critical revision of the manuscript for important intellectual content:* All authors.

*Statistical plan and analysis:* Faust, Du, Li, Lin

*Administrative, technical, or material support:* Mayes.

*Supervision:* Faust, Lin, Krumholz

## References

1. Holland KM, Jones C, Vivolo-Kantor AM, et al. Trends in US Emergency Department Visits for Mental Health, Overdose, and Violence Outcomes Before and During the COVID-19 Pandemic. JAMA Psychiatry. Published online February 3, 2021. doi:10.1001/jamapsychiatry.2020.4402

2. Federal Bureau of Investigations. Overview of Preliminary Uniform Crime Report, January–June, 2020 — FBI. Federal Bureau of Investigations. Accessed February 12, 2021. https://www.fbi.gov/news/pressrel/press-releases/overview-of-preliminary-uniform-crime-report-january-june-2020

3. National Highway Traffic Safety Administratio. Behavioral Research. NHTSA. Published December 19, 2016. Accessed February 12, 2021. https://www.nhtsa.gov/behavioral-research

4. National Center for Health Statistics O. Monthly Counts of Deaths by Select Causes, 2020-2021 |Data |Centers for Disease Control and Prevention. Accessed February 12, 2021. https://data.cdc.gov/NCHS/Monthly-Counts-of-Deaths-by-Select-Causes-2020-202/9dzk-mvmi

5. US Centers for Disease Control and Prevention. CDC WONDER. Accessed February 12, 2021. https://wonder.cdc.gov/

6. Huskamp HA, Busch AB, Uscher-Pines L, Barnett ML, Riedel L, Mehrotra A. Treatment of Opioid Use Disorder Among Commercially Insured Patients in the Context of the COVID-19 Pandemic. JAMA. 2020;324(23):2440. doi:10.1001/jama.2020.21512

